# Aeromedical retrieval diagnostic trends during a period of Coronavirus 2019 lockdown

**DOI:** 10.1101/2020.08.16.20176230

**Authors:** Fergus W Gardiner, Marianne Gillam, Leonid Churilov, Pritish Sharma, Mardi Steere, Michelle Hannan, Andrew Hooper, Frank Quinlan

## Abstract

**Background:** We aimed to compare the pre, lockdown, and post-lockdown aeromedical retrieval (AR) diagnostic reasons and patient demographics during a period of Coronavirus 2019 (COVID-19) social isolation.

**Methods:** An observational study with retrospective data collection, consisting of Australians who received an AR between 26 January and 23 June 2020. The main outcome measures were patient diagnostic category proportions and trends prior (28 January to 15 March), during (16 March to 4May), and following (5 May to 23 June 2020) social isolation restrictions.

**Results:** There were 16981 ARs consisting of 1959 (11.5) primary evacuations (PEs) and 12724 (88.5) inter-hospital transfers (IHTs), with a population median age of 52 years (interquartile range [IQR] 29.0–69.0), with 49.0% (n = 8283) of the cohort being male and 38.0% (n = 6399) being female. There were a total of six confirmed and 209 suspected cases of COVID-19, with the majority of cases (n = 114; 53.0%) in the social isolation period. As compared to pre-restriction, the odds of retrieval for the restriction and post-restriction period differed across time between the major diagnostic groups. This included, an increase in cardiovascular retrieval for both restriction and post-restriction periods(OR 1.12 95% CI 1.02-1.24 and OR 1.18 95% CI 1.08-1.30 respectively), increases in neoplasm in the post restriction period (OR 1.31 95% CI 1.04–1.64), and increases for congenital conditions in the restriction period (OR 2.56 95% CI 1.39-4.71). Cardiovascular and congenital conditions had increased rates of priority 1 patients in the restriction and post restriction periods. There was a decrease in endocrine and metabolic disease retrievals in the restriction period (OR 0.72 95% CI 0.53-0.98). There were lower odds during the post-restriction period for a retrievals of the respiratory system (OR 0.78 95% CI 0.67-0.93), and disease of the skin (OR 0.78 95% CI 0.6-1.0). Distribution between the 2019 and 2020 time periods differed (p< 0.05), with the lockdown period resulting in a significant reduction in activity.

**Conclusion:** The lockdown period resulted in increased AR rates of circulatory and congenital conditions. However, this period also resulted in a reduction of overall activity, possibly due to a reduction in other infectious disease rates, such as influenza, due to social distancing.

What is already known on this subject
We know the rates of Coronavirus 2019 (COVID-19) have an impact on a range of health services globally, however we do not have a clear understanding of the aeromedical retrieval reasons during a period of sustained social isolation.

What this study adds
While there were fewer aeromedical retrievals during the social isolation period there were relatively more retrievals for cardiovascular disease and congenital malformations of the circulatory system. However, post-restriction demonstrated reductions in respiratory disease and skin infections, as compared to the pre-restriction period. Many aeromedical retrievals come from areas with higher chronic disease rates. If these areas have community transmission of COVID-19, it is likely that retrievals will increase for severe COVID-19 patients, with associated comorbidities such as cardiovascular disease.

## Introduction

The novel coronavirus severe acute respiratory syndrome coronavirus 2 (SARS-CoV-2) originated in Wuhan China, and spread globally affecting millions of people. The major symptoms of coronavirus disease (COVID-19) include fever, cough, muscular soreness, and dyspnea,^1^ with most deaths occurring in older patients with at least one comorbidity.^2^ The current median reproductive number (R0) of COVID-19 is estimated by the World Health Organisation (WHO) to be 1.95, however recent studies indicate it could be closer to 2.79.^3^

The World Health Organisation (WHO) declared COVID-19 a pandemic on March 11 2020.^4^ COVID-19 rapidly spread throughout the world, with 118 319 cases reported on March 11 rising to more than 17 million by the end of July 2020.^5^ The rapid spread of COVID-19 led many countries to close their borders to international travel and impose social distancing and lockdown isolation measures, aimed at reducing or minimising community transmition.^4^

Lockdown measures differed slightly between geographic areas throughout Australia, however generally included: closures of non-essential business, working from home, national/state border closures, interpersonal social distancing, curfews, limitations on outdoor activities, school closures, personal hygiene measures, and recommendations associated with the use of personal protective equipment (PPE). Fines were issued for any major breaches.

The public health measures associated with controlling the virus, led to widespread economic disruptions,^6^ and potentially positive environmental impacts,^7,8^ such as reduced carbon emissions due to reduced travel. The global lockdown has also affected population health in other ways, such as reduced influenza transmission,^9^ reduced accidents and injuries,^10^ and potentially reduced preterm labour.^11^ Many of these reports are anecdotal observations, however it is likely that changes in the work environments, social interactions, and a focus of improved hand hygiene, has reduced overall exposure to other non-COVID infectious diseases.

To date there has been little research that considers whether nationwide lockdowns contribute to changes in patient diagnostic reasons, including those who require emergency aeromedical retrieval.

We aimed to investigate:

- whether the COVID-19 social lockdown/restriction was associated with differences in the rates and diagnostic reasons for people requiring aeromedical retrieval throughout Australia.
- whether the diagnostic proportions during the COVID-19 social lockdown/restriction period differed to patient diagnostic proportions seen in 2019.

## Methods

### Setting

During the COVID-19 pandemic the Royal Flying Doctor Service (RFDS) was tasked with transferring confirmed and suspected COVID-19 patients throughout Australia, in addition to its normal activity of 30,000-35,000 aeromedical transfers per year.^12^ Australian spans 7.69 million square kilometres, with the majority of the 2018 Australian population (n = 24,992,860) residing in major cities (n = 18,003,544; 72.0%) or inner-regional areas (n = 4,445,356; 17.8%), with the remainder living in outer-regional (n = 2,052,366; 8.2%), remote (n = 291,213; 1.2%), and very remote (n = 200,381; 0.1%) areas.^13^ With a fleet of 74 aircraft the RFDS provides extensive aeromedical retrieval for patients requiring treatment in inner-regional or major city hospitals.^14^

### Study Design

We conducted an observational study with retrospective data collection to investigate the diagnostic reasons patients were accessing aeromedical services throughout Australia. This analysis included all aeromedical retrievals conducted by the RFDS from 26 January to 23 June 2020. To determine whether this pandemic period differed to the same period in 2019, we included data from 26 January to 23 June 2019 to allow comparison.

An aeromedical retrieval was classified into two main categories, including primary evacuation (PE) (also known as primary retrieval) and inter-hospital transfer (IHT) (also known as a secondary retrieval). All repatriations were excluded from the analysis. A PE is generally staffed by a medical doctor and a nurse, while an IHT can only include a nurse or a medical doctor depending on the patient’s severity.

### Measures

Basic demographic information on age and gender was collected during the aeromedical retrieval. Aeromedical retrieval in-flight diagnoses were retrospectively coded to the International Statistical Classification of Diseases and Related Health Problems 10th Revision (ICD-10).^15^ During the aeromedical retrieval patients were categorised into priority categories from 1 to 3, with priority 1 being the most urgent.

To investigate the association between the time and differences in the rates and diagnostic reasons for people requiring aeromedical retrieval throughout Australia, each patient interaction was allocated to either pre-restriction, restriction/lockdown, and post-restriction time period depending on the date they received treatment in 2020. All three periods were chosen to last for 7-weeks and were defined as follows: pre-restriction chosen as the baseline period (28 January to 15 March); restriction from the time that Australia went into lockdown (16 March to 4 May) and post-restriction where restrictions were gradually eased (5 May to 23 June).

The outcome measures included the number of patients across the three study periods (each 7-weeks duration), the proportions and percentages for the ICD-10 diagnoses and patient demographics, across the three periods studied.

### Statistical analysis

Continuous variables were summarised as medians and interquartile ranges (IQRs). Categorical variables were summarised as counts and proportions. The association between the time period and the likelihood of requiring aeromedical retrieval due to a specific diagnostic reason in 2020 was investigated using logistic regression with magnitudes of association reported as odds ratios (ORs) with respective 95% confidence intervals (95% CIs) using pre-restriction period as a reference category. No p-values for individual diagnostic categories are reported due to the number of categories investigated. 95% CIs that do not include 1 are indicative of statistical significance at two-sided threshold of 0.05. The frequency distribution of the air ambulance retrievals across the three studies were compared between 2019 and 2020, using Chi-square test. Statistical analyses were performed using Stata 15IC statistical software (StataCorp, College Station, TX, USA) with graphs implemented in the statistical software package R version 3.5.1.

### Ethics approval

This project was deemed a low-risk quality assurance project. As this project involved routinely collected data, specific patient consent forms were not required.

### Patient and Public Involvement

This study was developed and informed by patient priorities. The RFDS is an essential service, with the RFDS being the only healthcare service provider in many areas. This research is broadly aimed at understanding patient trends to promote and provide evidence-based services to the Australian public. RFDS stakeholders, including patients, will receive study results through various formats, including social media, newsletters, and via community engagement. Social engagement will also be used to determine community needs associated with COVID-19 lockdowns, especially with Aboriginal and Torres Strait Islander (Indigenous) communities. This will help to ensure our pandemic responses are culturally appropriate.

## Results

There were 16981 aeromedical retrievals throughout the 2020 study period including 1959 (11.5%) PEs, and 15022 IHTs (88.5%), as compared to 18125 retrievals consisting of 2337 (12.9%) PEs and 15788 (87.1%) IHTs during the same date range in the year 2019.

The 2019 activity consistently increased across the 3 time periods from 5793 (32.0%), 6140 (33.9%), to 6192 (34.2%). However, in the pandemic study period there were observed differences in the distribution, in terms of the number of retrievals, with 5927 (34.9%), 5121 (30.2%), and 5933 (34.9%), prior, during, and following social isolation restrictions, respectively. The 2020 pandemic period resulted in a reduction in overall activity, as compared to the same time period in 2019. Historical trends were expected to increase with the Australian winter months (June, July, August), however the lockdown period resulted in a reduction in activity as compared to the same timeframe in the prior year (see Figure 2).

**Figure 1:**
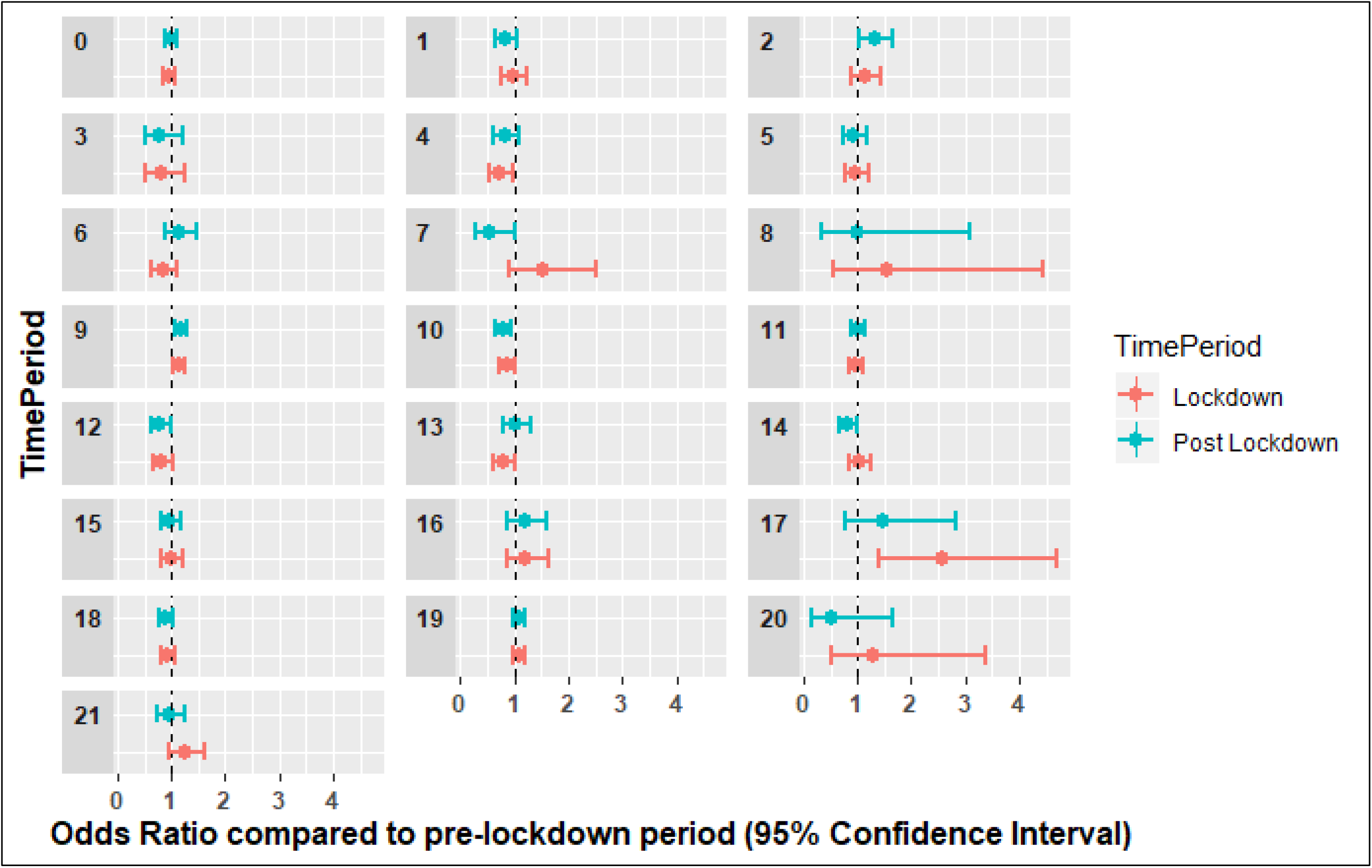
Odds ratio forest plot by ICD-10 chapter for the lockdown and post-lockdown periods.

**Figure 2:**
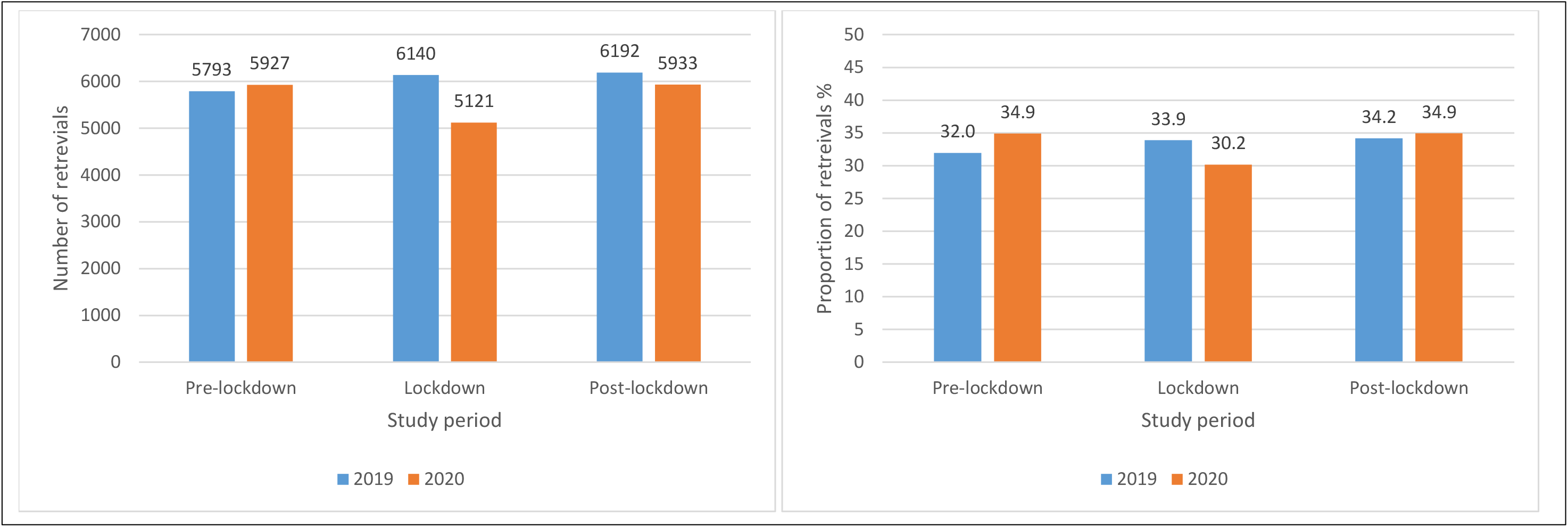
The year 2020 Pandemic period compared to the same timeframe in the year 2019.

The median population age of patients that underwent an aeromedical retrieval by the RFDS during the 2020 pandemic study period was 52 years (interquartile range [IQR] 29.0–69.0). There were 8283 (49.0%) males, 6399 (38.0%) females, and 2299 (13.5%) missing gender, with the male median age being 55 (IQR:55-70) and the female median age being 47 (IQR: 26-67) years, with the female median age reflecting retrievals that were related to pregnancy (n = 639; 3.8%).

The leading retrieval reasons throughout the study period included diseases of the circulatory system (n = 3082; 18.15%), injury (n = 2768; 16.3%), and diseases of the digestive system (n = 1535; 9.0%). There were significantly lower numbers of aeromedical retrievals performed during the period of social isolation (n = 5121; p< 0.05), compared to the pre-isolation period (n = 5927). See Table 1 for diagnostic trends.

**Table 1:**
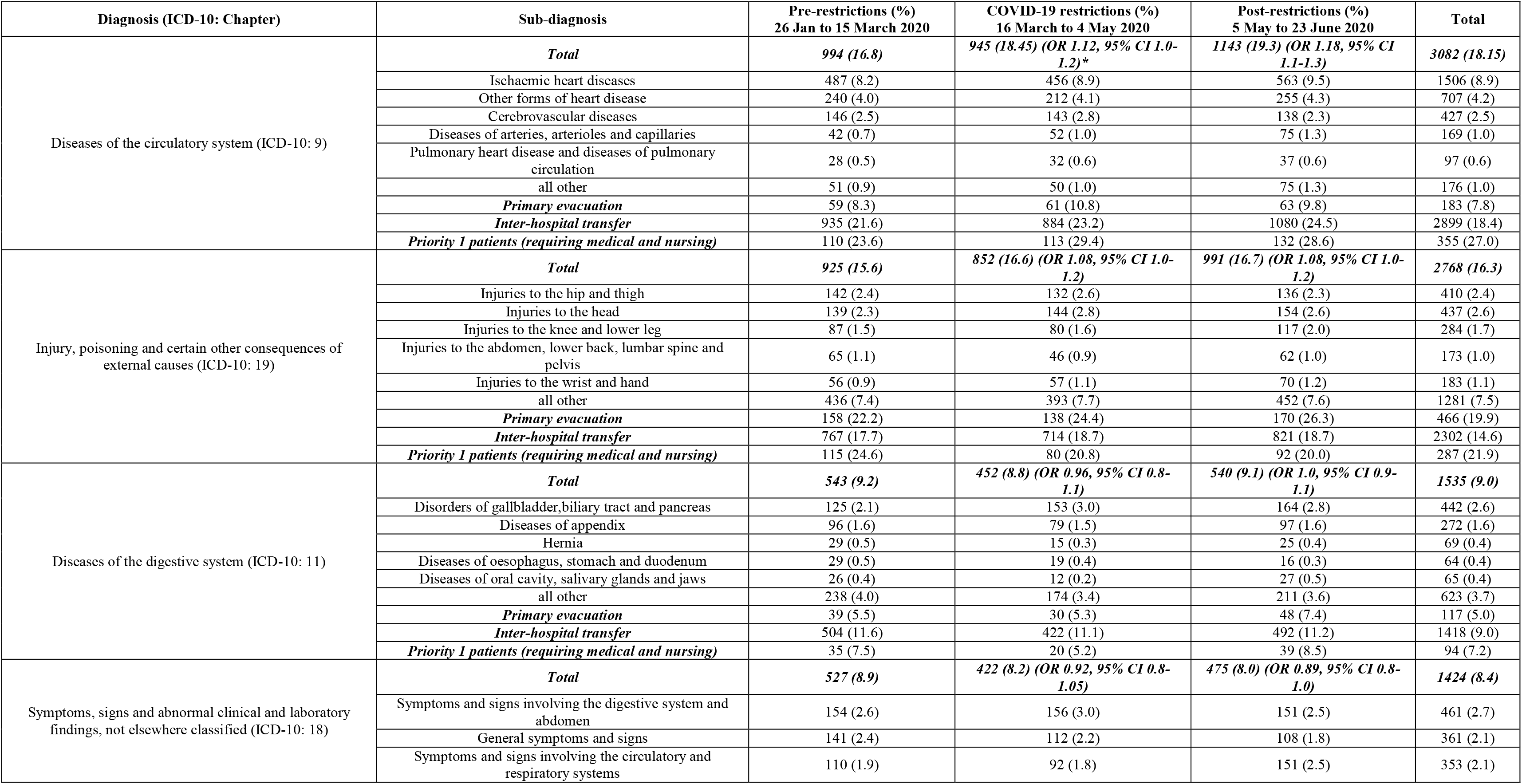
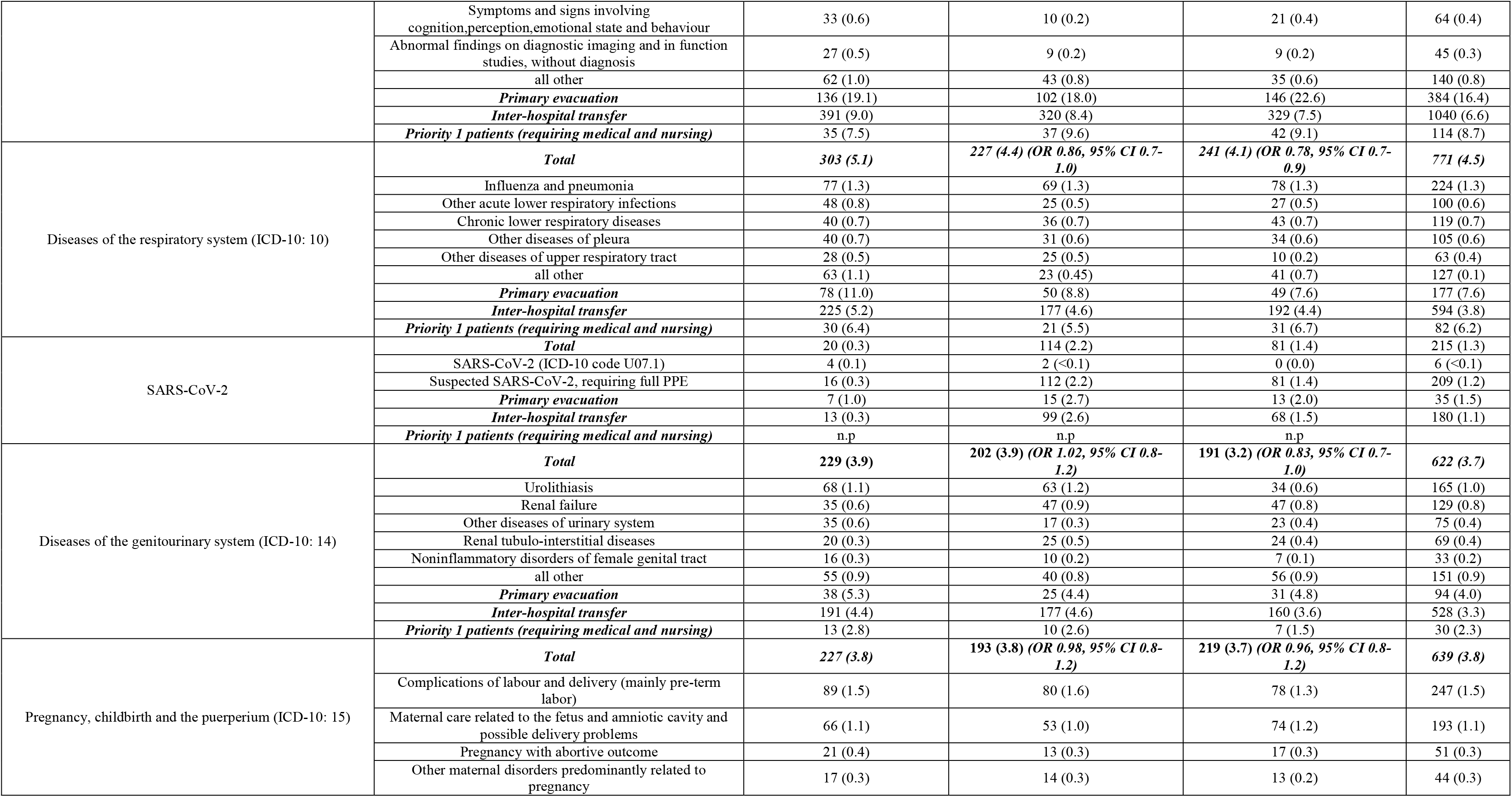
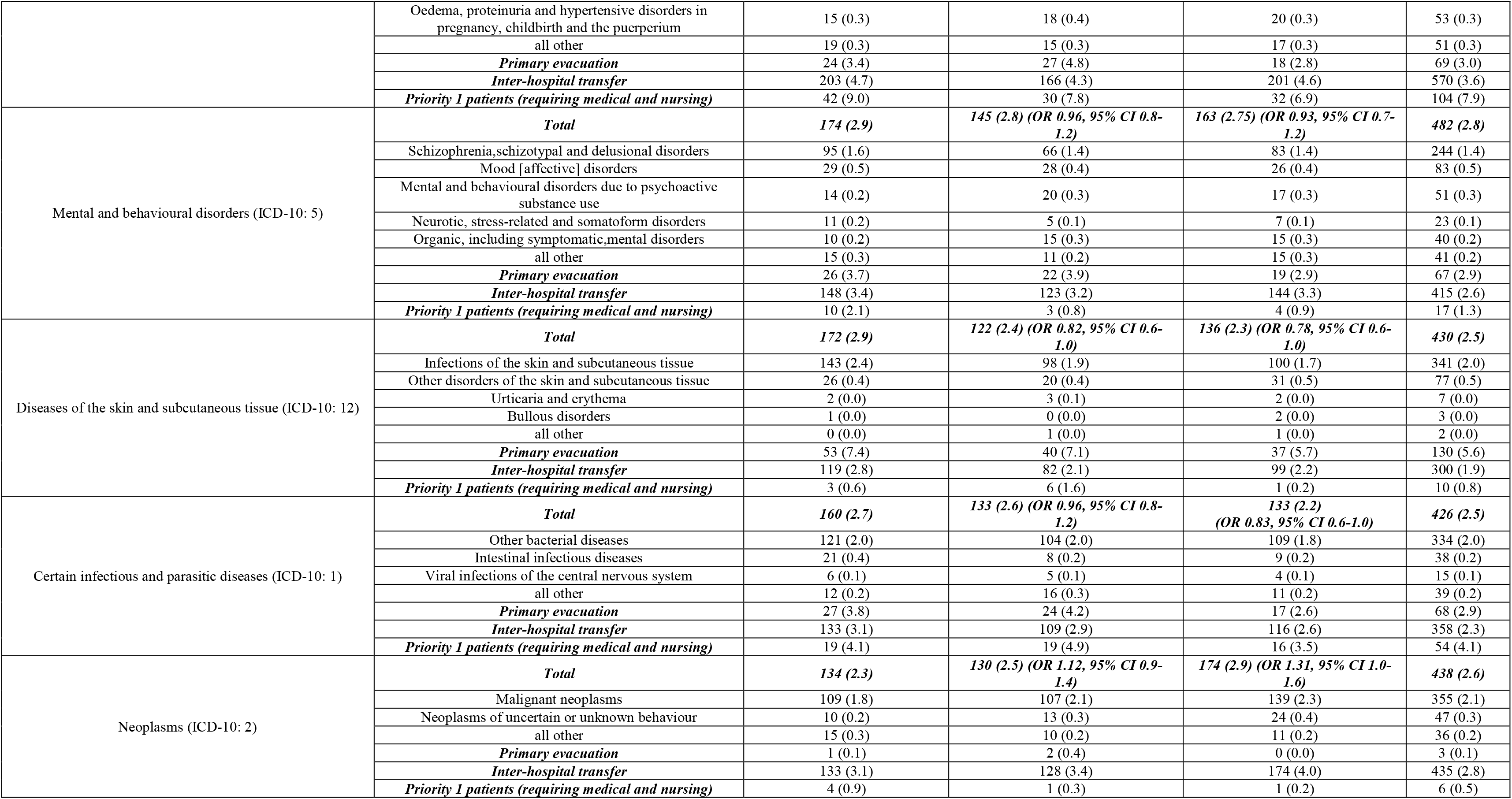
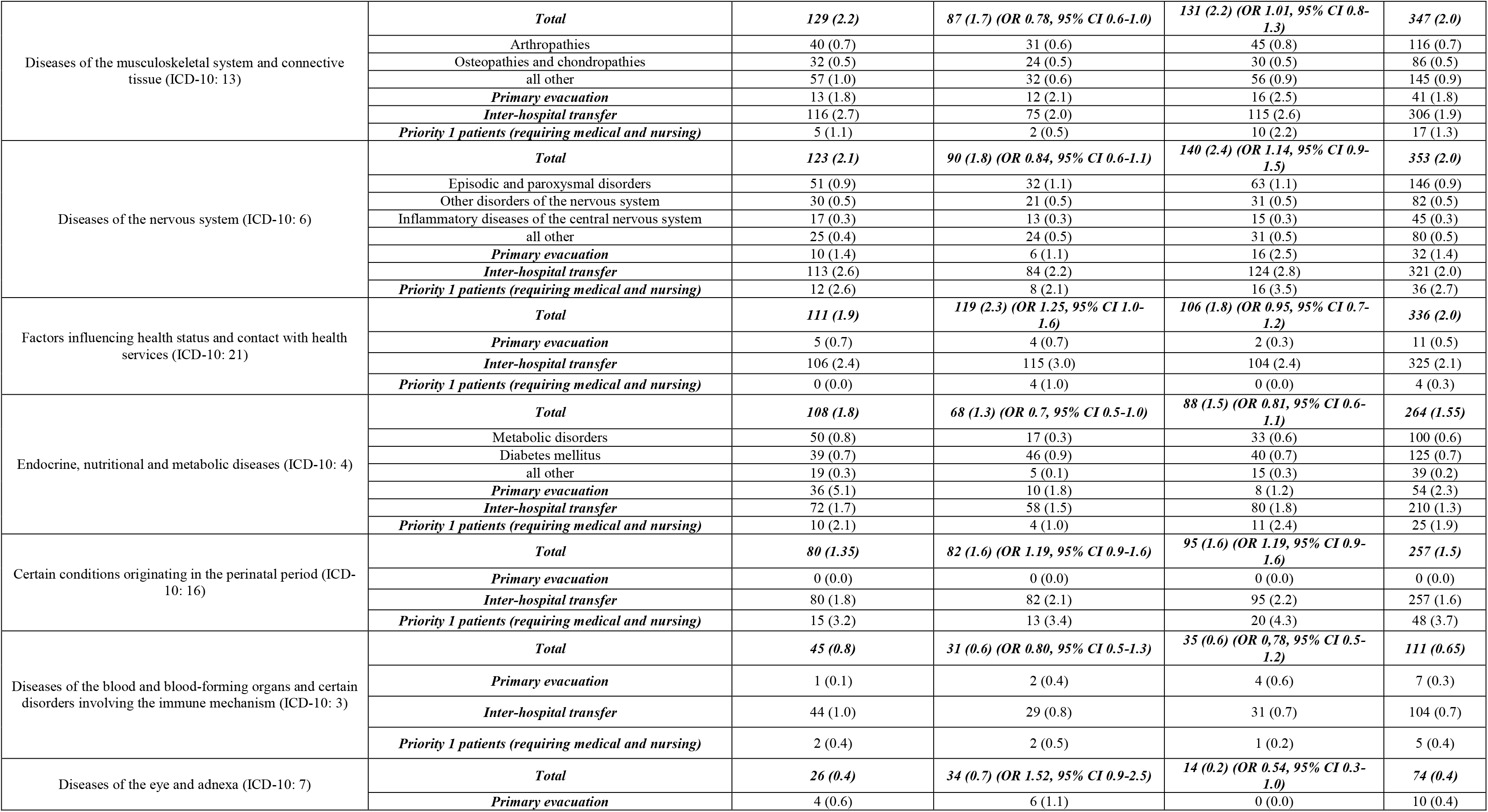
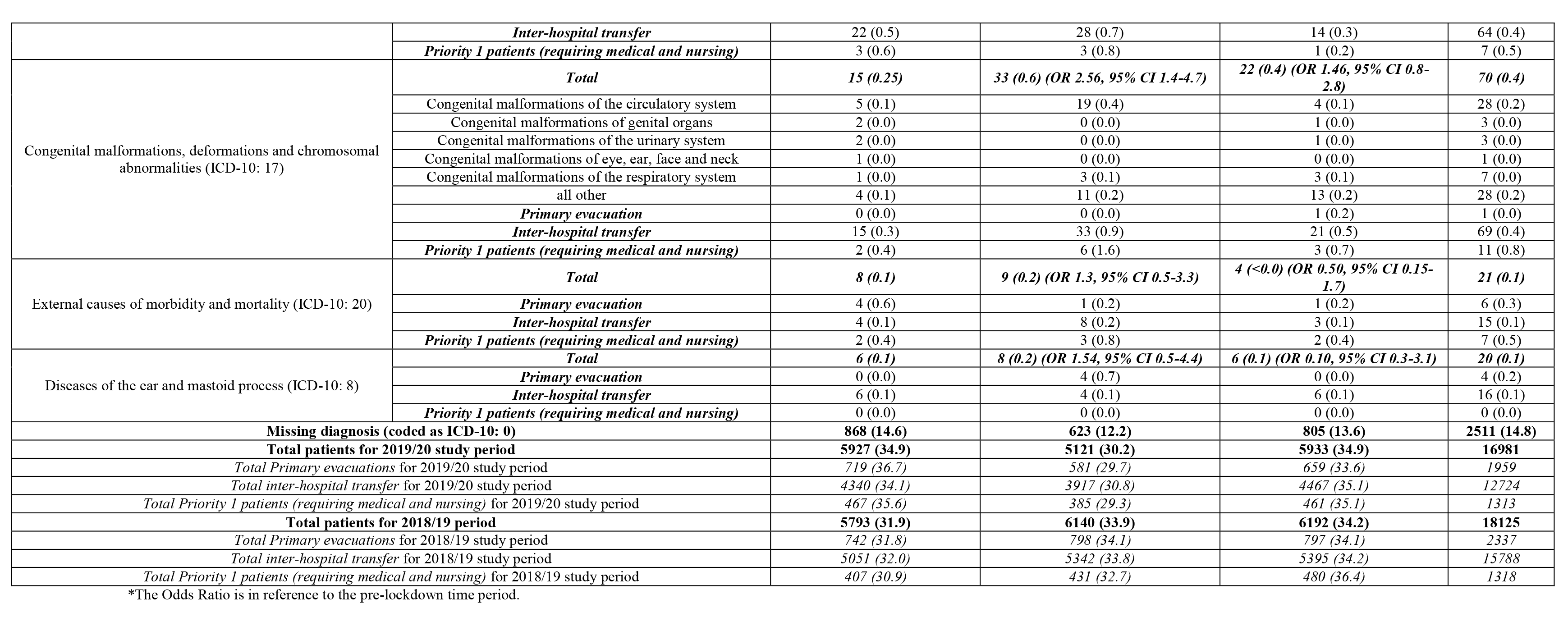
Aeromedical retrieval diagnosis trends prior, during, and post a period of COVID-19 restrictions.

There were six confirmed and 209 aeromedical COVID-19 cases, with the majority (n = 114; 53.0%) conducted during the social isolation period. It should be noted, that all the areas that these patients came from did not pathology testing services. As such all suspected cases were treated as confirmed cases.

Cardiovascular retrievals decreased in overall number during lockdown (n = 945; 18.45%) however started to increase in number during the post-lockdown period (n = 1143;19.3%). Compared to the pre-restriction period there was an increase in odds of cardiovascular retrievals during the lockdown and post-restriction periods (OR 1.12 95% CI 1.02-1.24 and OR 1.18 95% CI 1.08-1.30 respectively), indicating a relative increase compared to other reasons for retrievals. The most common diagnosis in this group was ischaemic heart disease (n = 1506; 8.9%). The restriction (n = 113; 29.4%) and post-restriction (n = 132; 28.6%) periods also had higher rates of cardiovascular priority 1 patients, as compared to the pre-restriction period (n = 110; 23.6%).

There was a decrease in number of retrievals for respiratory diseases during restriction and an increase in number during the post-restriction period. There were significantly lower odds of retrieval for respiratory diseases in the post-restriction period compared to pre-restriction, indicating a relative decrease of respiratory system retrievals compared to other reasons (OR 0.78 95% CI 0.67-0.93). The restriction (n = 21; 5.5%) period had lower rates, while the post-restriction period had higher rates (n = 31; 6.7%) of respiratory priority 1 patients, as compared to the pre-restriction period (n = 30; 6.4%).

The number of retrievals for diseases of the skin and subcutaneous tissue were in the restriction period compared to the pre-restriction period, however in the post restriction period the number was higher than in the two prior periods as were the odds of retrieval (OR 0.78 95% CI 0.62-0.98) indicating a relative increase of retrievals in the post-restriction period. The number of retrievals for neoplasm were similar in the restriction period compared to the pre-restriction period, however in the post restriction period the number was higher than in the two prior periods as were the odds of retrieval (OR 1.31 95% CI 1.04-1.64) indicating a relative increase of neoplasm retrievals in the post-restriction period.

There were lower number of retrievals both in the restriction and the post-restriction period for the endocrine and metabolic diagnostic groups. The odds of retrieval for this diagnostic group in the restriction period was significantly lower than the odds in the pre-restriction period (OR 0.72 95% CI 0.53-0.98), indicating relatively less retrievals for this diagnosis in the restriction period compared to the pre-restriction period.

The number of retrievals for congenital conditions diagnosis was low across the three periods, however higher in the restriction and post-restriction period than in the pre-restriction period. The odds of retrievals for congenital conditions during the restriction period were significantly higher than in the pre-restriction period (OR 2.56 95% CI 1.39-4.71). The restriction (n = 6; 1.6%) and post-restriction (n = 3; 0.7%) period also had slightly higher rates of congenital priority 1 patients, as compared to the pre-restriction period (n = 2; 0.4%).

## Discussion

As an essential service, the RFDS provides emergency aeromedical support throughout Australia, as reflected in recent COVID-19 activity reports.^16^ The main findings from this paper indicated that there were relatively more retrievals for diseases of the circulatory system, and congenital malformations, deformations and chromosomal abnormalities, during the social isolation period compared to the pre-isolation period. During the post-isolation period there was an increase in number of retrievals for diseases of the circulatory system and for neoplasms, however a reduction for respiratory disease, and skin conditions. The increases of cardiovascular and neoplasm treatment during this period may indicate a rebound effect in patients subsequently seeking treatment following social isolation. Furthermore, the decrease in respiratory and skin infections during the post-lockdown period may indicated that social distancing is also influencing other disease transmission rates, such as influenza.

Many of the retrievals for the circulatory system were for ischaemic heart disease, such as acute myocardial infarction and angina pectoris. It is unclear why there were relatively less circulatory cases however a higher proportion during the lockdown period. This may reflect patients delaying early invention due to social isolation measures and limitations in accessing primary healthcare services.^17,18^ This appeared to be the case, as there were more circulatory system cases during the post-lockdown period.

Cardiovascular disease, older adults, those with respiratory disease, and diabetes are at the highest risk of complications from COVID-19 infection. While many of our patients would not be considered old, with a median age of 52 years old, many of the retrievals were for endocrine disease, and specifically uncontrolled diabetes. Many of these patients come from rural and remote areas, which also have high rates of metabolic syndrome.^18^ Uncontrolled diabetes, or chronic hyperglycaemia, negatively affects the immune system increasing morbidity and mortality due to increased infection risk and organic complications.^19^ For example, those with Influenza A (H1N1) and diabetes, have triple the risk of hospitalization and quadruple the risk of intensive care unit (ICU) admission once hospitalized.^20^ It is likely that COVID-19 infection coupled with diabetes would have similar rates,^20^ as the mortality rates in China indicate that 42.3% of their COVID-19 deaths had diabetes.^21^

A striking finding was that there were significantly more paediatric patients retrieved for congenital malformations of the circulatory system during the lockdown period, most requiring a IHT from a rural area to a large metropolitan hospital mainly for cardiac surgery and congenital heart defects. This was due to limitations with accessing local cardiac surgeons/ units. There is limited data that details the effects that COVID-19 has on the paediatric population, due to less than 1% of hospitalised cases to date being children younger than 10 years old.^22^ While children seem less vulnerable to severe COVID-19,^23^ those with congenital malformations are likely to have worse COVID-19 outcomes.^23^

During the lockdown period there were less aeromedical PE and IHT transfers for cancer, increasing significantly post-lockdown. This trend is believed to be related to many oncologists within Australia only conducting essential treatments. The lockdown period provided many challenges associated with cancer patient safety, specifically that patients would need to leave their homes and thus would expose themselves to infection.^24^ Cancer treatment in itself predisposes patients to the more serious effects of COVID-19. For example, a recent study found that patients with cancer are at a greater risk of death and/or intensive care unit admission (OR 5.4, 95% CI 1.8–16.2).^25^ Oncologists were required to weigh the risks of death and morbidity from COVID-19 against the benefits of cancer therapy. This risk is arguably amplified during aeromedical retrieval and transferring patients hundreds of kilometres from their homes into inner-regional and major city areas. The risks associated with transporting patients for cancer treatment are believed to be the main driver for a reduction in transfers during the lockdown period.

It was found that the retrievals for diseases of the respiratory system started to decline during the isolation period, and significantly reduced in the post-isolation period. This was mainly driven by reductions in acute respiratory disease retrievals. While influenza retrieval rates did not appear to reduce, reductions in overall respiratory disease match findings that social distancing for COVID-19 is reducing other respiratory disease transmission rates. For example, the Australian rates of confirmed influenza has significantly reduced since increased hygiene and social isolation measures have been in place, as compared to the winter months of 2019.^9^ However, this trend could also be a result of a strong influenza vaccination campaign operated by the Australian Government during the early isolation periods. As per respiratory disease, skin infection retrieval rates started to decrease during the lockdown period, and significantly reduced in the post-lockdown period. As observed within Australia and internationally, non-pharmaceutical interventions, including border restrictions, quarantine and isolation, distancing, and changes in population behaviour, not only reduce COVID-19 transmission but also led to reductions in other diseases.^26^

The vast majority of aeromedical retrievals come from rural and remote areas of Australia.^13^ These areas are characterised as having healthcare workforce shortages, coupled with higher social and economic disadvantage and higher rates of chronic disease, including cardiovascular and respiratory disease.^27^ A potential contributor to the higher relative rates of retrievals for cardiovascular disease could be associated with limitations in these population groups able to access telehealth services, as traditional face-to-face services during the lockdown period were limited. As many remote Australian communities have limited access to COVID-19 testing and management facilities, coupled with higher chronic disease risk factors, it is likely that during mass infection many of these patients will require air ambulance transfer to inner-regional and major city hospitals.^16^

This study highlighted interesting trends during a period of social isolation and lockdown. Future research should consider whether the overall numerical reductions in cardiovascular disease were due to less clinical cases, or a result of less people seeking treatment. Attention to the higher rates of congenital malformations of the circulatory system should also be considered in the context of future paediatric research. Future research should also consider social isolation trends within primary healthcare.

This study was limited to one medical service; however, this limitation was reduced by the RFDS being the largest aeromedical service in the world, providing services to the whole of the Australian continent during the study period. This study did not include primary healthcare interactions prior to aeromedical retrieval, however this limitation will be addressed in a future study.

## Conclusion

Social isolation measures led to a reduction in overall aeromedical activity, however increased significantly once the restrictions were lifted. During the social isolation period there were relatively more retrievals for cardiovascular disease, such as ischaemic heart disease, and congenital malformations of the circulatory system compared to pre-restriction. These population groups if infected with COVID-19 have significantly worse outcomes, and should be considered a high risk population group. Many of the Australian aeromedical retrievals come from rural and remote areas, characterised by vast geographical distances, often with significantly lower healthcare provision. If these community areas have community transmission of COVID-19, it is likely that aeromedical retrievals will increase for severe COVID-19 patients, with associated comorbidities such as cardiovascular disease.

## Data Availability

Upon request.

## Notes

### Competing Interest Statement

The authors have declared no competing interest.

### Funding Statement

No external funding was received

### Author Declarations

This project was deemed a low-risk quality assurance project by the RFDS Clinical and Health Services Research Committee (CHSRC), which provides oversight for RFDS research projects. As this project involved routinely collected data, specific patient consent forms were not required.

